# Deep phenotyping of 34,128 patients hospitalised with COVID-19 and a comparison with 81,596 influenza patients in America, Europe and Asia: an international network study

**DOI:** 10.1101/2020.04.22.20074336

**Authors:** Edward Burn, Seng Chan You, Anthony G. Sena, Kristin Kostka, Hamed Abedtash, Maria Tereza F. Abrahão, Amanda Alberga, Heba Alghoul, Osaid Alser, Thamir M Alshammari, Maria Aragon, Carlos Areia, Juan M. Banda, Jaehyeong Cho, Aedin C Culhane, Alexander Davydov, Frank J DeFalco, Talita Duarte-Salles, Scott DuVall, Thomas Falconer, Sergio Fernandez-Bertolin, Weihua Gao, Asieh Golozar, Jill Hardin, George Hripcsak, Vojtech Huser, Hokyun Jeon, Yonghua Jing, Chi Young Jung, Benjamin Skov Kaas-Hansen, Denys Kaduk, Seamus Kent, Yeesuk Kim, Spyros Kolovos, Jennifer C.E. Lane, Hyejin Lee, Kristine E Lynch, Rupa Makadia, Michael E. Matheny, Paras P. Mehta, Daniel R Morales, Karthik Natarajan, Fredrik Nyberg, Anna Ostropolets, Rae Woong Park, Jimyung Park, Jose D. Posada, Albert Prats-Uribe, Gowtham Rao, Christian Reich, Yeunsook Rho, Peter Rijnbeek, Lisa M. Schilling, Martijn Schuemie, Nigam H. Shah, Azza Shoaibi, Seokyoung Song, Matthew Spotnitz, Marc A. Suchard, Joel N. Swerdel, David Vizcaya, Salvatore Volpe, Haini Wen, Andrew E. Williams, Belay B. Yimer, Lin Zhang, Oleg Zhuk, Daniel Prieto-Alhambra, Patrick Ryan

**Author notes:** Corresponding author Corresponding author: Professor Daniel Prieto-Alhambra, Centre for Statistics in Medicine, NDORMS, University of Oxford. Joint first authors (Edward Burn, Seng Chan You).

## Abstract

**Background:** In this study we phenotyped individuals hospitalised with coronavirus disease 2019 (COVID-19) in depth, summarising entire medical histories, including medications, as captured in routinely collected data drawn from databases across three continents. We then compared individuals hospitalised with COVID-19 to those previously hospitalised with influenza.

**Methods:** We report demographics, previously recorded conditions and medication use of patients hospitalised with COVID-19 in the US (Columbia University Irving Medical Center [CUIMC], Premier Healthcare Database [PHD], UCHealth System Health Data Compass Database [UC HDC], and the Department of Veterans Affairs [VA OMOP]), in South Korea (Health Insurance Review & Assessment [HIRA]), and Spain (The Information System for Research in Primary Care [SIDIAP] and HM Hospitales [HM]). These patients were then compared with patients hospitalised with influenza in 2014-19.

**Results:** 34,128 (US: 8,362, South Korea: 7,341, Spain: 18,425) individuals hospitalised with COVID-19 were included. Between 4,811 (HM) and 11,643 (CUIMC) unique aggregate characteristics were extracted per patient, with all summarised in an accompanying interactive website (http://evidence.ohdsi.org/Covid19CharacterizationHospitalization/). Patients were majority male in the US (CUIMC: 52%, PHD: 52%, UC HDC: 54%, VA OMOP: 94%,) and Spain (SIDIAP: 54%, HM: 60%), but were predominantly female in South Korea (HIRA: 60%). Age profiles varied across data sources. Prevalence of asthma ranged from 4% to 15%, diabetes from 13% to 43%, and hypertensive disorder from 24% to 70% across data sources. Between 14% and 33% were taking drugs acting on the renin-angiotensin system in the 30 days prior to hospitalisation. Compared to 81,596 individuals hospitalised with influenza in 2014-19, patients admitted with COVID-19 were more typically male, younger, and healthier, with fewer comorbidities and lower medication use.

**Conclusions:** We provide a detailed characterisation of patients hospitalised with COVID-19. Protecting groups known to be vulnerable to influenza is a useful starting point to minimize the number of hospital admissions needed for COVID-19. However, such strategies will also likely need to be broadened so as to reflect the particular characteristics of individuals hospitalised with COVID-19.

## Introduction

The ongoing coronavirus disease 2019 (COVID-19) pandemic is placing a huge strain on health systems worldwide. While a number of studies have provided information on the clinical characteristics of individuals being hospitalised with COVID-19,[1–3] substantial uncertainty around the prevalence of comorbidities and prior medication use among this population remains. Moreover, it is not known whether those hospitalised with COVID-19 are systematically different to individuals hospitalised during previous influenza seasons. Providing such information would help to inform the current response to COVID-19.

COVID-19 shares similarities with influenza to the extent that both cause respiratory disease which can vary markedly in its severity and present with a similar constellation of symptoms, including fever, cough, myalgia, malaise and fatigue, and dyspnea. Early reports do, however, indicate that the proportion of severe infections and mortality rate is higher for COVID-19.[4] Older age and a range of underlying health conditions, such as immune deficiency, cardiovascular disease, chronic lung disease, neuromuscular disease, neurological disease, chronic renal disease, and metabolic diseases, have been associated with an increased risk of severe influenza and associated mortality.[5] While age appears to be a clear risk factor for severe COVID-19,[4] other associations are not yet well understood. Comparisons with COVID-19 are further complicated by the heterogeneity in influenza itself, with different strains resulting in different clinical presentations and associated risks. Those hospitalised with the A(H1N1)pdm09 subtype of the influenza A virus during the associated influenza pandemic in 2009 were, for example, generally younger and with fewer comorbidities than those from preceding influenza seasons.[6]

Routinely-collected health care data can improve our understanding of the characteristics of individuals hospitalised with COVID-19, with years of prior clinical observations recorded. In this study, our first aim was to characterise in detail the demographics and medical histories of individuals hospitalised with COVID-19 across multiple institutions in three countries from North America (United States), Europe (Spain) and Asia (South Korea). Subsequently, we aimed to compare those hospitalised with COVID-19 with patients hospitalised with influenza in previous seasons.

## Methods

### Study design

This is a cohort study based on routinely-collected primary care and hospital electronic health records (EHRs), hospital billing data, and insurance claims data from the US, South Korea, and Spain. The data sources used were mapped to the Observational Medical Outcomes Partnership (OMOP) Common Data Model (CDM).[7] The open-science Observational Health Data Sciences and Informatics (OHDSI) network maintains the OMOP CDM, and its members have developed a wide range of tools to facilitate analyses of such mapped data.[8] Two particular benefits of this approach were that contributing centres did not need to share patient-level data and common analytical code could be applied across databases.

### Data sources

Data from the US, South Korea, and Spain underpinned the study. EHR data from the US came from the Columbia University Irving Medical Center (CUIMC), covering NewYork-Presbyterian Hospital/Columbia University Irving Medical Center, University of Colorado Health Data Compass (UC HDC), which includes the UCHealth System with data from 12 hospitals, and United States Department of Veterans Affairs (VA OMOP), which includes 170 medical centers. In addition, data from a US hospital billing system database came from the Premier Hospital database (PHD). EHR data from Spain came from The Information System for Research in Primary Care (SIDIAP), a primary care records database that covers approximately 80% of the population of Catalonia, Spain,[9] and the inpatient care database of HM Hospitales (HM), a hospital group which includes fifteen general hospitals from all over Spain, with detailed hospital admission information for COVID-19 patients from March 1^st^ to April 20^th^ 2020. Data from South Korea came from Health Insurance Review & Assessment (HIRA), a repository of national claims data which is collected in the process of reimbursing healthcare providers.[10] In addition, the analysis was also performed on US EHR data from Tufts-Clinical Academic Research Enterprise Trust (CLARET), which covers Tufts Medical Center (TMC), and the STAnford medicine Research data Repository (STARR-OMOP), which includes data from Stanford Health Care,[11] and on data from the Daegu Catholic University Medical Center, a teaching hospital in Daegu, South Korea, covered by Federated E-health Big Data for Evidence Renovation Network (FEEDER-NET). These latter sites are not reported here due to smaller study populations, but the results are reported in the accompanying interactive website (http://evidence.ohdsi.org/Covid19CharacterizationHospitalization/).

### Study participants

Patients hospitalised between December 2019 and April 2020 with COVID-19 were identified on the basis of having a hospitalisation along with a confirmatory diagnosis or test result of COVID-19 within a time window from 21 days prior to admission up to the end of their hospitalisation. This time window was chosen so as to include those who had the diagnosis made prior to their hospitalisation and allow for a delay in test results or diagnoses to be made or recorded. Patients were also required to be aged 18 years or older at the time of hospitalisation. The same algorithm was used to identify COVID-19 cases across sites, except for CUIMC where the algorithm was adjusted to account for local coding practice.

Analogous criteria were used for identifying individuals hospitalised with influenza between September 2014 to April 2019, with individuals identified on the basis of having a hospitalisation along with a confirmatory diagnosis or test result of influenza within a time window from 21 days prior to admission up to the end of their hospitalisation. For SIDIAP, influenza cases were only available up to the end of 2017. An additional cohort of those hospitalised with influenza between September 2009 and April 2010 was also identified. The motivation for this latter group was that the 2009-2010 flu epidemic included many cases of A(H1N1)pdm09 infection, which had different clinical characteristics and associated severity compared to the seasonal flu. Each individual’s first hospitalisation with COVID-19 or a particular flu season was considered.

Except for the HM and Premier databases, individuals were required to have a minimum of 365 days of prior observation time available for the primary analysis, to allow for a comprehensive capture of baseline diagnoses and medications prior to their hospitalisation. As this restriction could exclude persons with little prior health care utilisation or without sustained health insurance, we also characterised cohorts without this requirement in a sensitivity analysis. Given the nature of data collection, individuals in HM and Premier had no prior observation time available and so no requirement for prior observation time was imposed.

### Patient features and data analysis

Age at hospitalisation and sex distributions were summarised. Medication use was calculated over three time periods: 1) from a year prior up to, and including, the day of hospitalisation, 2) from 30 days prior up to, and including, the day of hospitalisation, and 3) the day of hospitalisation. Only the latter time period was reported for HM and Premier. Drug eras were calculated to give the span of time when an individual is assumed to be exposed to a particular active ingredient. These begin on the start date of the first drug exposure and end on the observed end date if available, or were inferred (for example, based on the number of days of supply). A persistence window of up to 30 days was permitted between two medication records for them to be considered as part of the same drug era. Individual medications were categorised using Anatomical Therapeutic Chemical (ATC) groupings. All drugs are reported in full in a dedicated interactive website (see Results section), but specific classes are reported here based on recent interest due to their potential effects (positive or negative) on COVID-19 susceptibility or severity: agents acting on the renin-angiotensin system (including angiotensin converting enzyme inhibitors (ACE) inhibitors and angiotensin II receptor blockers (ARBs)), antiepileptics, beta blocking agents, calcium channel blockers, diuretics, drugs for acid related disorders, immunosuppressants, insulins and analogues, and lipid modifying agents (such as statins). Prevalence of medication use for each time window was determined by the proportion of persons who had at least one day during the time window overlapping with a drug era for each medication or drug class of interest. Conditions were identified on the basis of SNOMED codes, with all descendent codes included. Similarly, all recorded diagnoses are available for consultation in the accompanying website, but a list of key conditions is reported here based on recent reports of associations with COVID-19 infection or outcomes.

Age distributions in each cohort were plotted. The proportion of a cohort having a particular characteristic was described, with the prevalence of all medications and conditions captured in the databases depicted using Manhattan style plots. Standardised mean differences (SMD) were calculated when comparing characteristics of study cohorts, with the SMD for all variables plotted in Manhattan style plots. In addition, the prevalence of particular conditions or medications of interest among those hospitalised with COVID-19 (Y axis) were compared to those hospitalised with influenza (X axis) in scatter plots, with dots on the top-left indicating a higher prevalence among those hospitalised with COVID-19 and dots on the bottom-right indicating a higher prevalence among those hospitalised with influenza. The analytic code used in the study has been made available at https://github.com/ohdsi-studies/Covid19HospitalizationCharacterization.

## Results

### Patients hospitalised with COVID-19

A total of 34,128 individuals hospitalised with COVID-19 (CUIMC: 1,759; HIRA: 7,341; HM: 2,078; PHD: 5,257; SIDIAP: 16,347; UC HDC: 769; VA OMOP: 577) were included. 68,829 summary characteristics, from 4,811 (HM) to 11,643 (CUIMC) unique aggregate characteristics, were extracted and summarised. All are made available in an accompanying interactive website (http://evidence.ohdsi.org/Covid19CharacterizationHospitalization/).

Cohorts from CUIMC, HM, PHD, SIDIAP, UC HDC, and VA OMOP were predominantly male (52%, 60%, 52%, 54%, 54%, and 94% respectively, but mostly female for HIRA (60%). The age distributions of those hospitalised for COVID-19 are summarised in Figure 1 (alongside those hospitalised with influenza, see below). Different patterns are seen in the various contributing databases with, in particular, patients in South Korea (HIRA) seen to be younger than elsewhere.

**Figure 1:**
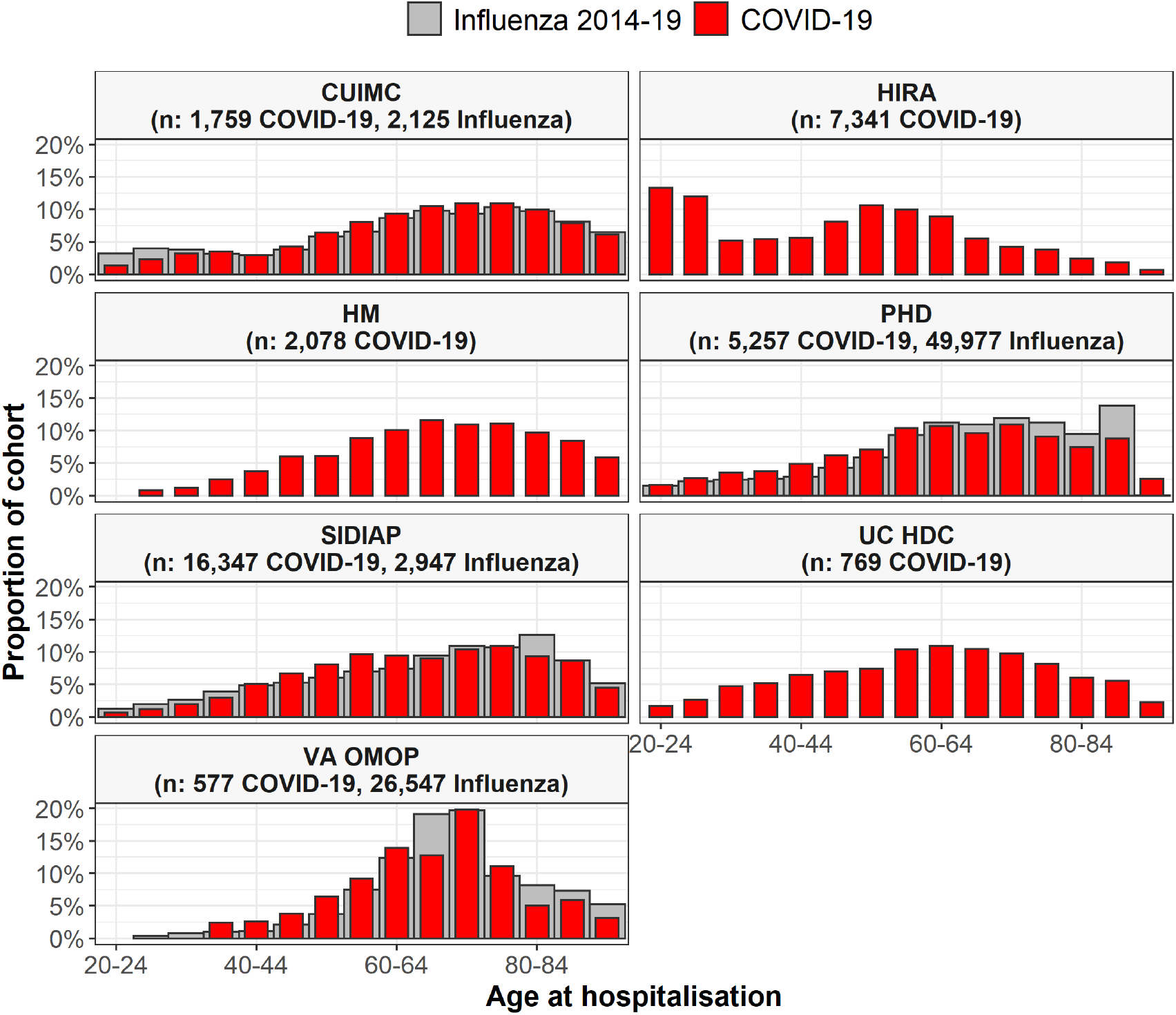
Age of patients hospitalised with COVID-19 and of patients hospitalised with influenza Individuals hospitalised with COVID-19 between December 2019 and April 2020 compared with those hospitalised with influenza between September 2014 to April 2019 (where available). Proportion of cohorts by 5-year age groups, with groups with counts of less than 10 omitted. CUIMC: Columbia University Irving Medical Center; HIRA: Health Insurance Review & Assessment; HM: HM Hospitales; PHD: Premier Healthcare Database; SIDIAP: The Information System for Research in Primary Care; UC HDC: University of Colorado Health Data Compass; VA OMOP: Department of Veterans Affairs. Influenza data for SIDIAP was only available from 2014 to 2017.

The distribution of comorbidities in COVID-19 patients varied across sites and countries, see Table 1. The mean Charlson comorbidity index of those hospitalised with COVID-19 in the US ranged from 3.1 for PHD to 5.4 for VA OMOP, from 0.8 for HM to 1.4 for SIDIAP in Spain, and was 2.7 in HIRA, covering South Korea. In the US, the proportion of those hospitalised with COVID-19 who had asthma ranged from 7% to 15%, from 24% to 43% for diabetes, from 28% to 49% for heart disease, and from 8% to 18% for cancer. In Spain, between 4% and 7% for asthma, from 13% to 22% for diabetes, from 17% to 27% for heart disease, and from 5% to 16% for cancer. In South Korea, 12% of those hospitalised had a history of asthma, 17% had diabetes, 13% heart disease, and 4% cancer. The prevalence of hypertension ranged from 37% to 70% in the US, from 30% to 46% in Spain, and was 24% in South Korea. The prevalence of the full range of conditions summarised are shown in Figure 2, with all values reported at http://evidence.ohdsi.org/Covid19CharacterizationHospitalization/. For medications, the proportion of those hospitalised with COVID-19 in the US who had been taking agents acting on the renin-angiotensin system over the 30 days prior to their hospitalisation ranged from 18% to 39%, while the proportions taking immunosuppressants ranged from 4% to 6%, and from 21% to 51% for lipid modifying agents over the same time period. In HIRA, 14% had been taking agents acting on the renin-angiotensin system, 1% immunosuppressants, and 16% lipid modifying agents. In SIDIAP, 27% had been taking agents acting on the renin-angiotensin system, 2% immunosuppressants, and 24% lipid modifying agents (Table 2). Looking at one medication of particular interest (of which many can be explored in detail at http://evidence.ohdsi.org/Covid19CharacterizationHospitalization/), use of hydroxychloroquine on the day of admission was 7% in HIRA and 41% in HM. The prevalence of the many medications summarised are shown in Figure 2, with all values reported http://evidence.ohdsi.org/Covid19CharacterizationHospitalization/. Removing the requirement of having a year prior history in datasets other than HM and Premier did not materially change the results (see http://evidence.ohdsi.org/Covid19CharacterizationHospitalization/ for full details).

**Table 1:** Conditions of individuals hospitalised with COVID-19

|  | CUIMC (n: 1,759) | HIRA (n: 7,341) | HM (n: 2,078) | PHD (n: 5,257) | SIDIAP (n: 16,347) | UC HDC (n: 769) | VA OMOP (n: 577) |
| --- | --- | --- | --- | --- | --- | --- | --- |
| Charlson index | 4.0 | 2.0 | 0.8 | 3.1 | 1.5 | 3.5 | 5.4 |
| Anemia | 13.1% | 11.9% | 4.2% | 30.2% | 14.6% | 29.9% | 30.0% |
| Anxiety disorder | 5.5% | 11.5% | 2.3% | 18.0% | 23.7% | 18.2% | 30.3% |
| Asthma | 7.2% | 12.0% | 4.2% | 14.6% | 6.9% | 13.8% | 9.9% |
| Atrial fibrillation | 8.4% | 1.0% | 6.8% | 18.1% | 8.2% | 12.0% | 15.8% |
| Chronic liver disease | 2.6% | 5.0% | 0.6% | 3.0% | 1.9% | 2.3% | 6.8% |
| COPD | 6.1% | 1.9% | 5.9% | 24.5% | 7.6% | 12.6% | 28.6% |
| Dementia | 8.5% | 5.3% | 2.3% | 9.2% | 5.6% | 7.4% | 7.8% |
| Diabetes mellitus | 24.2% | 16.6% | 13.2% | 35.8% | 21.6% | 35.2% | 43.3% |
| GERD | 9.6% | 32.3% | 1.6% | 24.8% | 9.5% | 23.4% | 26.9% |
| Heart disease | 28.3% | 13.1% | 16.5% | 48.6% | 27.1% | 39.0% | 48.2% |
| Heart failure | 10.7% | 5.1% | 2.7% | 24.8% | 5.9% | 12.9% | 22.2% |
| Hyperlipidemia | 24.7% | 31.1% | 21.3% | 46.0% | 23.7% | 38.0% | 55.8% |
| Hypertensive disorder | 36.6% | 23.8% | 29.6% | 47.1% | 45.6% | 57.3% | 69.7% |
| Insomnia | 3.1% | 4.2% | 0.7% | 4.4% | 14.6% | 7.4% | 10.9% |
| Ischemic heart disease | 8.0% | 6.9% | 4.1% | 16.0% | 7.3% | 13.5% | 15.6% |
| Low back pain | 6.8% | 23.2% | <0.5% | 5.0% | 21.9% | 11.1% | 30.0% |
| Malignant neoplastic disease | 7.7% | 4.4% | 4.5% | 9.4% | 15.8% | 10.8% | 18.4% |
| Osteoarthritis of knee | 4.6% | 7.7% | <0.5% | 2.0% | 14.8% | 5.1% | 11.4% |
| Osteoarthritis of hip | 3.0% | 0.5% | <0.5% | 1.1% | 5.1% | 2.6% | 2.8% |
| Peripheral vascular disease | 4.7% | 8.0% | 0.5% | 7.5% | 3.9% | 7.4% | 10.6% |
| Renal impairment | 21.1% | 1.8% | 7.6% | 37.5% | 13.1% | 37.2% | 32.9% |
| Venous thrombosis | 2.3% | 0.6% | 0.8% | 2.0% | 1.5% | 6.6% | 4.0% |
| Viral hepatitis | 2.3% | 3.9% | 0.6% | 2.5% | 1.7% | 2.6% | 5.9% |

CUIMC: Columbia University Irving Medical Center; HIRA: Health Insurance Review & Assessment; HM: HM Hospitales; PHD: Premier Healthcare Database; SIDIAP: The Information System for Research in Primary Care; UC HDC: University of Colorado Health Data Compass; VA OMOP: Department of Veterans Affairs. COPD: Chronic obstructive pulmonary disease; GERD: Gastroesophageal reflux disease. Exact proportions have not been reported where counts were less than 10. Conditions were identified over the year prior to hospitalisation (note, HM and PHD had no prior time requirement with conditions primarily recorded on day of hospital admission).

**Table 2:** Prior medications of individuals hospitalised with COVID-19

|  | CUIMC (n:<br>1,759) | HIRA (n:<br>7,341) | HM (n:<br>2,078) | PHD (n:<br>5,257) | SIDIAP (n:<br>16,347) | UC HDC (n:<br>769) | VA OMOP (n:<br>577) |
| --- | --- | --- | --- | --- | --- | --- | --- |
| Antineoplastic and immunomodulating agents |  |  |  |  |  |  |  |
| Year prior to hospitalisation | 9.6 % | 4.5 % | - | - | 3.4 % | 12.9% | 13.2% |
| 30 days prior to hospitalisation | 6.1 % | 2.6 % | - | - | 2.7 % | 9.6% | 7.1% |
| At hospitalisation | 4.9 % | 2.0 % | 3.8 % | 2.1 % | 2.5 % | 8.2% | 4.9% |
| Agents acting on the renin-angiotensin system |  |  |  |  |  |  |  |
| Year prior to hospitalisation | 27.3% | 15.9% | - | - | 30.2% | 39.8% | 49.6% |
| 30 days prior to hospitalisation | 18.0% | 14.2% | - | - | 26.6% | 33.0% | 38.5% |
| At hospitalisation | 15.9% | 13.4% | 9.9% | 7.4% | 25.7% | 31.5% | 33.1% |
| Antiepileptics |  |  |  |  |  |  |  |
| Year prior to hospitalisation | 19.0 % | 9.1 % | - | - | 10.6 % | 27.2% | 33.4% |
| 30 days prior to hospitalisation | 9.9 % | 5.2 % | - | - | 8.1 % | 21.6% | 22.7% |
| At hospitalisation | 9.1 % | 4.2 % | 3.3 % | 11.3 % | 7.7 % | 20.2% | 16.6% |
| Anti-inflammatory and antirheumatic products |  |  |  |  |  |  |  |
| Year prior to hospitalisation | 23.6 % | 63.9 % | - | - | 30.9 % | 52.9% | 46.1% |
| 30 days prior to hospitalisation | 10.5 % | 30.0 % | - | - | 9.9 % | 38.6% | 16.8% |
| At hospitalisation | 7.8 % | 17.7 % | 3.8 % | 7.9 % | 6.3 % | 35.1% | 9.9% |
| Antithrombotic agents |  |  |  |  |  |  |  |
| Year prior to hospitalisation | 44.9% | 34.6% | - | - | 25.7% | 72.0% | 46.1% |
| 30 days prior to hospitalisation | 29.9% | 17.1% | - | - | 21.9% | 67.5% | 16.8% |
| At hospitalisation | 28.3% | 11.4% | 38.2% | 38.4% | 21.3% | 66.8% | 9.9% |
| Beta blocking agents |  |  |  |  |  |  |  |
| Year prior to hospitalisation | 26.9 % | 10.1 % | - | - | 13.8 % | 32.8% | 44.4% |
| 30 days prior to hospitalisation | 16.8 % | 6.5 % | - | - | 12.4 % | 24.7% | 36.0% |
| At hospitalisation | 15.2 % | 5.6 % | 5.7 % | 17.8 % | 11.9 % | 23.8% | 31.0% |
| Calcium channel blockers |  |  |  |  |  |  |  |
| Year prior to hospitalisation | 23.5 % | 16.7 % | - | - | 12.4 % | 24.3% | 35.0% |
| 30 days prior to hospitalisation | 15.0 % | 14.6 % | - | - | 10.5 % | 20.5% | 27.0% |
| At hospitalisation | 13.4 % | 14.0 % | 6.8 % | 9.7 % | 10.1 % | 19.9% | 24.3% |
| Diuretics |  |  |  |  |  |  |  |
| Year prior to hospitalisation | 27.6 % | 10.2 % | - | - | 24.6 % | 33.3% | 42.6% |
| 30 days prior to hospitalisation | 19.2 % | 7.5 % | - | - | 21.5 % | 27.4% | 33.1% |
| At hospitalisation | 17.7 % | 6.7 % | 7.6 % | 14.1 % | 20.7 % | 26.4% | 28.4% |
| Drugs for acid related disorders |  |  |  |  |  |  |  |
| Year prior to hospitalisation | 35.4 % | 68.8 % | - | - | 30.4 % | 53.6% | 52.0% |
| 30 days prior to hospitalisation | 20.6 % | 36.5 % | - | - | 22.7 % | 44.5% | 36.2% |
| At hospitalisation | 17.9 % | 29.3 % | 39.7 % | 21.8 % | 21.5 % | 42.4% | 28.9% |
| Immunosuppressants |  |  |  |  |  |  |  |
| Year prior to hospitalisation | 5.6 % | 2.0 % | - | - | 2.3 % | 7.5% | 5.7% |
| 30 days prior to hospitalisation | 3.9 % | 0.7 % | - | - | 1.9 % | 6.1% | 3.5% |
| At hospitalisation | 3.7 % | 0.4 % | 2.5 % | 1.5 % | 1.7 % | 5.9% | 2.6% |
| Insulins and analogues |  |  |  |  |  |  |  |
| Year prior to hospitalisation | 17.8% | 3.7% | - | - | 5.5% | 31.9% | 22.9% |
| 30 days prior to hospitalisation | 10.9% | 2.7% | - | - | 4.9% | 26.9% | 18.4% |
| At hospitalisation | 9.9% | 2.4% | <0.5% | 13.1% | 4.7% | 25.4% | 14.6% |
| Lipid modifying agents |  |  |  |  |  |  |  |
| Year prior to hospitalisation | 33.8% | 18.4% | - | - | 26.8% | 41.7% | 61.9% |
| 30 days prior to hospitalisation | 20.8% | 15.5% | - | - | 23.6% | 36.4% | 50.8% |
| At hospitalisation | 18.7% | 14.1% | 5.4% | 19.9% | 23.0% | 35.6% | 43.8% |

CUIMC: Columbia University Irving Medical Center; HIRA: Health Insurance Review & Assessment; HM: HM Hospitales; PHD: Premier Healthcare Database; SIDIAP: The Information System for Research in Primary Care; UC HDC: University of Colorado Health Data Compass; VA OMOP: Department of Veterans Affairs. Study participants from HM and PHD were not required to have a year of prior history and so only medications at time of hospitalisation are summarised.

**Figure 2:**
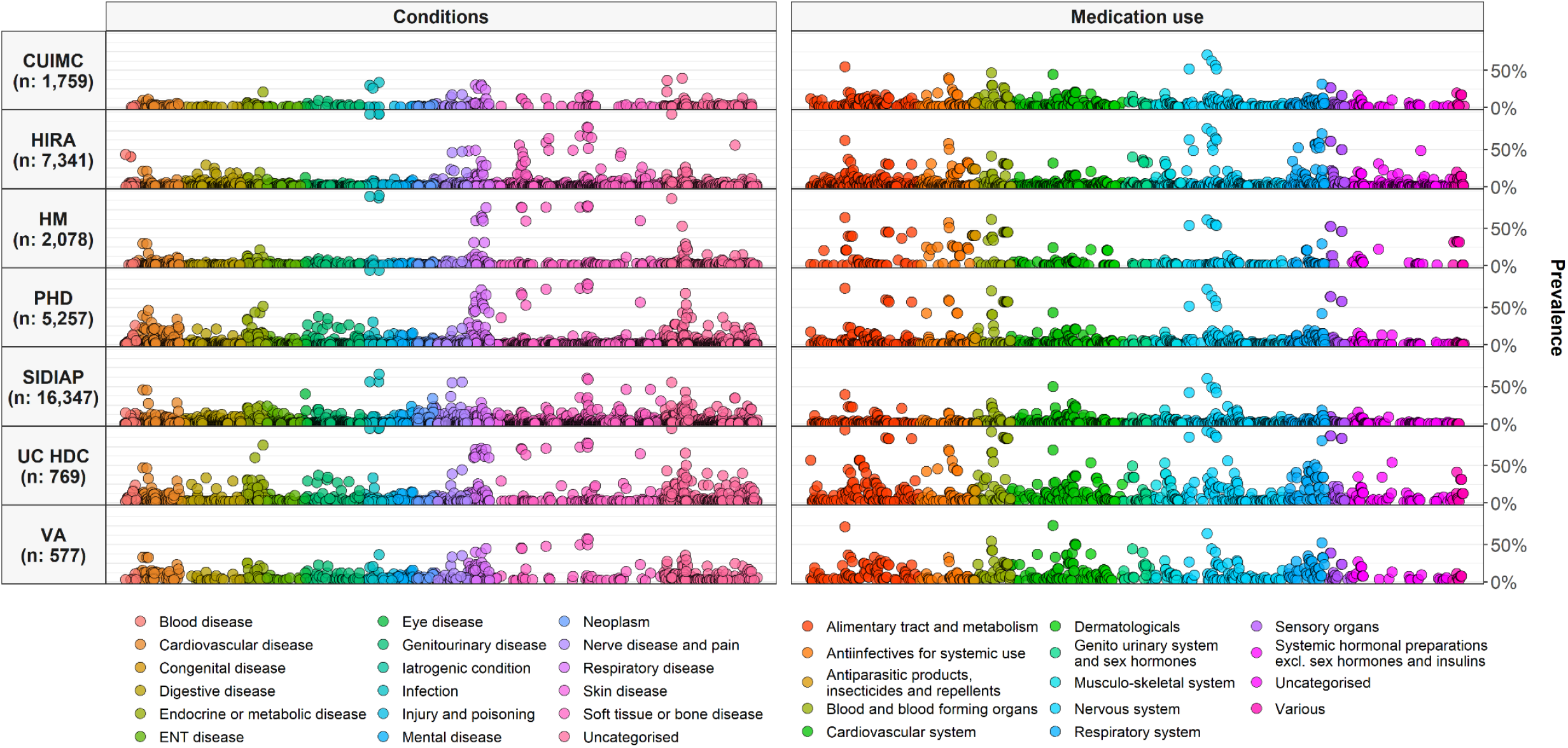
Prevalence of conditions and medication use among COVID-19 patients. Individuals hospitalised with COVID-19 between December 2019 and April 2020. Conditions from up to a year prior, medication use from day of hospitalisation. CUIMC: Columbia University Irving Medical Center; HIRA: Health Insurance Review & Assessment; HM: HM Hospitales; PHD: Premier Healthcare Database; SIDIAP: The Information System for Research in Primary Care; UC HDC: University of Colorado Health Data Compass; VA OMOP: Department of Veterans Affairs.

### A comparison of patients hospitalised with COVID-19 and patients hospitalised with influenza

A total of 81,596 patients hospitalised with influenza between 2014 to 2019 (2,125 CUIMC, 49,977 PHD, 2,947 SIDIAP, 26,547 VA OMOP). In addition, 2,443 patients hospitalised with influenza between 2009 to 2010 were included (170 CUIMC, 1,689 SIDIAP, 584 VA OMOP) were also identified. Patient characteristics of those hospitalised with COVID-19 are compared to those of individuals hospitalised with influenza between 2014 and 2019 in Figures 1, 3, and 4, and with those hospitalised with influenza between 2009 and 2010 in Appendix Figures 1 and 2.

**Figure 3:**
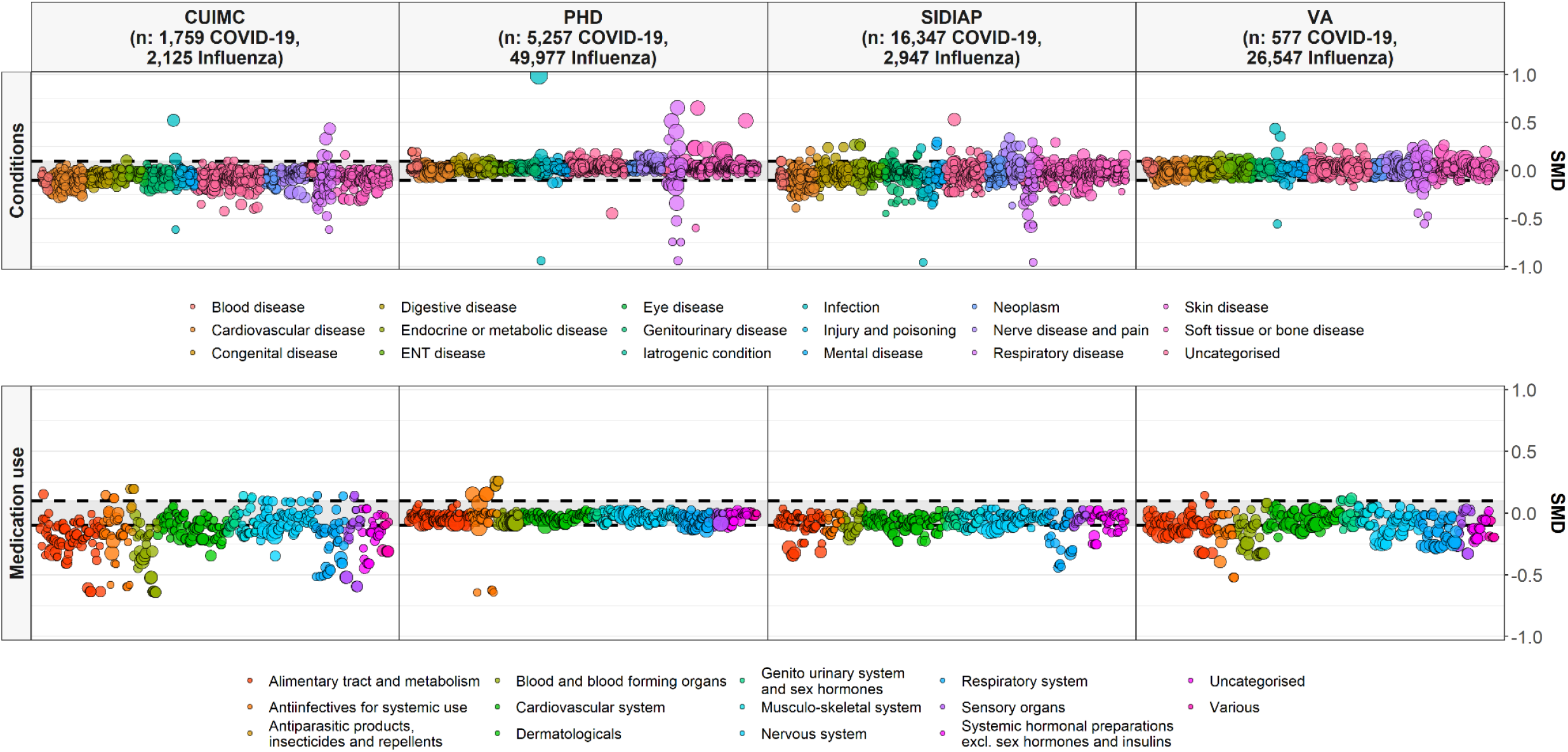
Standardised mean difference in conditions (top) and medication use (bottom) among COVID-19 patients compared to 2014-2019 influenza patients Individuals hospitalised with COVID-19 between December 2019 and April 2020 compared with those hospitalised with influenza between September 2014 to April 2019. Conditions from up to a year prior, medication use from day of hospitalisation. Each dot represents one of these covariates with the colour indicating the absolute value of the standardised mean difference (SMD), with a SMD above 0.1 taken to indicate a difference in the prevalence of a particular covariate, and with the size of the dot reflecting the prevalence of the variable in the COVID-19 study populations. CUIMC: Columbia University Irving Medical Center; PHD: Premier Healthcare Database; SIDIAP: The Information System for Research in Primary Care; VA OMOP: Department of Veterans Affairs.

**Figure 4:**
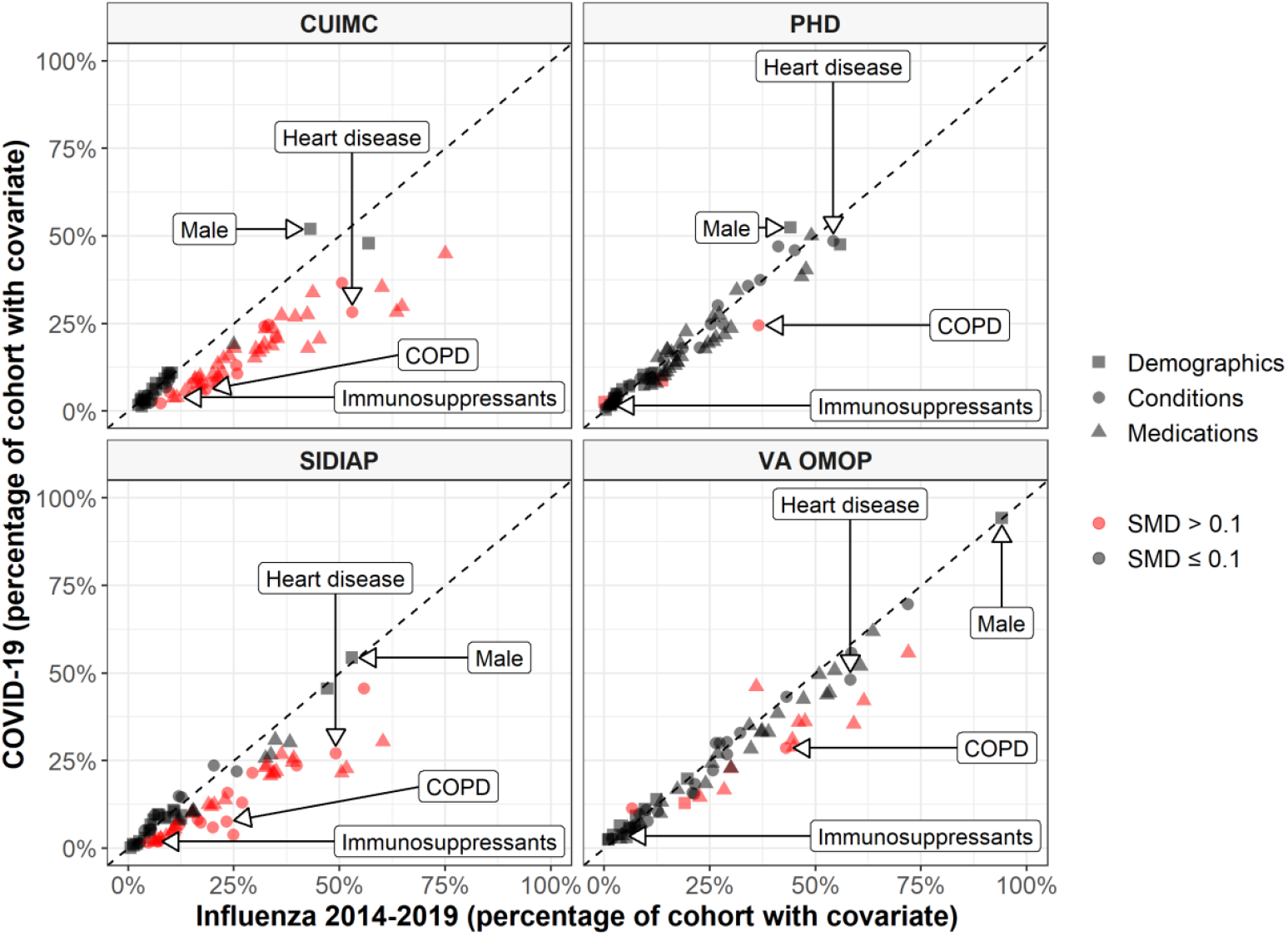
Characteristics of COVID-19 patients compared to 2014-2019 Influenza patients The plot compares demographics (age and sex), conditions (recorded over the year prior and up to day of hospitalisation), and medications (1) from a year prior to day of hospitalisation, 2) from 30 days prior to day of hospitalisation, and 3) on day of hospitalisation). Each dot represents one of these covariates with the colour indicating the absolute value of the standardised mean difference (SMD), with a SMD above 0.1 taken to indicate a difference in the prevalence of a particular covariate. The proportion male, with heart disease, with chronic obstructive pulmonary disease (COPD), and taking immunosuppressants (over the 30 days prior to hospitalisation) are shown for illustration. CUIMC: Columbia University Irving Medical Center; PHD: Premier Healthcare Database; SIDIAP: The Information System for Research in Primary Care; VA OMOP: Department of Veterans Affairs.

A greater proportion of those hospitalised with COVID-19 were male compared to those hospitalised with influenza between 2014 to 2019 for CUIMC, PHD, and SIDIAP. Of those hospitalised between 2014 to 2019 with influenza 43%, 44%, 53% were male for each of these respective data sources. The ages of those hospitalised with COVID-19 generally appeared slightly younger compared to those hospitalised with influenza between 2014 to 2019, see Figure 1.

Those hospitalised with COVID-19 had a comparable or lower prevalence of comorbidities compared to those hospitalised with influenza 2014-2019 (see Figure 3 top and Figure 4). Chronic obstructive pulmonary disease (COPD), cardiovascular disease and dementia were all more common amongst those hospitalised with influenza compared to those hospitalised with COVID-19. Medication use was less common amongst COVID-19 patients (Figure 3 bottom and Figure 4) with, for example, systemic corticosteroids and alpha-blockers amongst those more frequently used among those hospitalised with influenza.

Those hospitalised with influenza between 2009 to 2010 were typically younger compared to both COVID-19 and to influenza 2014-2019 admissions (see Appendix Figure 1, and Appendix Table 2).

COVID-19 patients were more likely to be male, with 40%, 49% and 91% of those hospitalised between 2009 to 2010 for influenza being male for CUIMC, SIDIAP, and VA OMOP. Comparisons of conditions and medications, however, varied depending on the data source (see Appendix Figure 4 and Appendix Table 2).

## Discussion

### Summary of key findings

The characteristics of 34,128 patients hospitalised with COVID-19 in US, South Korea, and Spain have been extracted from EHRs and health claims databases and summarised. Between 5,000 and 12,000 unique aggregate characteristics have been produced across databases, with all made publicly available in an accompanying website.

Patients hospitalised with COVID-19 in the US and Spain were predominantly male with age distributions varying across data sources, while those in South Korea were mostly female and appreciably younger than patients in the US and Spain. Many comorbidities were common among individuals hospitalised with COVID-19 with, as an example, 37% to 70% of those hospitalised with COVID-19 in the US, 30% to 46% of those in Spain, and 24% of those in South Korea having hypertension. Similarly, prior medication use was common with, for example, 18% to 39% in the US, 27% in Spain, and 14% in South Korea, taking drugs acting on the renin-angiotensin system (ACE inhibitors and ARBs) in the 30 days prior to their hospitalisation.

Comparisons with previous cohorts of patients admitted to hospital for seasonal influenza in recent years suggest that COVID-19-related admissions are seen more often in younger patients and with a higher proportion men. In the US and Spain, those hospitalised with COVID-19 were generally either of comparable health or healthier than patients hospitalised with influenza. Consistent differences were noted in the prevalence of respiratory disease, cardiovascular disease and dementia, each more common amongst those hospitalised with influenza in all of the contributing databases. Similarly, the use of corticosteroids and alpha-blockers was consistently higher amongst influenza patients. Those hospitalised with influenza in 2009-2010, during the pandemic associated with H1N1, were seen to be younger than both those hospitalised with influenza in more recent years and patients hospitalised with COVID-19.

### Findings in context

A number of studies have previously provided information on individuals hospitalised with COVID-19. While cohorts have generally been majority male, the prevalence of comorbidities have varied. In a study of 1,099 individuals who tested positive for COVID-19 in China, of whom 94% were hospitalised, 58% were male, with 7% having diabetes, 15% hypertension, and 1% cancer.[12] In another study of 191 patients with COVID-19 in two hospitals in Wuhan, China, 62% were male, 19% had diabetes, 30% had hypertension, and 1% had cancer.[13] In a study which identified 1,999 individuals who tested positive for COVID-19 and were hospitalised in New York, 63% were male, 25% had diabetes, 10% COPD, and 45% a cardiovascular condition.[14] In another US study of 1,482 patients admitted to hospital with COVID-19 in March 2020, 55% were male, with 28% having diabetes, 11% having COPD, and 28% having cardiovascular disease.[15] Our findings add to this emerging body of evidence. The results from our study also provide an illustration of the variation in patient characteristics across contexts, with heterogeneity seen both across the cohorts from the US and between the US, Spain, and South Korea.

The comparison with influenza made in our study adds important context when considering the characteristics of those hospitalised with COVID-19. Individuals hospitalised with COVID-19 appear to be more likely younger and male and, in the US and Spain, to have fewer comorbidities than those hospitalised with influenza in previous years. Indeed, those hospitalised with COVID-19 were consistently seen to be less likely to have COPD, cardiovascular disease and dementia than those hospitalised with influenza in recent years.

This study has also added important information on medication use by individuals hospitalised with COVID-19, based on electronic health records and claims data. There is tremendous interest in the risks and benefits of medications such as ACE-inhibitors and ARBs for COVID-19, and whether other medications, such as ibuprofen, should be avoided. However, to date, there has been little evidence as to what proportion of those hospitalised with COVID-19 have previously been taking such medications. Our findings shed light on this area, and highlight the importance of further research on the benefits and harms associated with continued use of such treatments, especially those that are commonly taken amongst individuals with COVID-19. It has been seen here, for example, that between 1 in 10 and 2 in 5 of those hospitalised with COVID-19 were taking medicines acting on the renin-angiotensin system in the month before their hospital admission. The consequences of temporarily discontinuing such treatments on cardiovascular risks and mortality remain unknown [16]. Interestingly, corticosteroid use, recently shown to be effective to treat COVID-19,[17] was consistently seen to be less prevalent in patients hospitalised with COVID-19 compared to those hospitalised with influenza across databases, as were alpha-blockers which some have hypothesised to have a beneficial effect in COVID-19.[18]

It should be noted that the characteristics of individuals with COVID-19 have been described in this study at the particular point in time of admission to hospital. While this is of particular interest given its intrinsic link with health care utilisation, this only provides a snapshot of the whole picture. Those testing positive for COVID-19 in the community without progressing to hospitalisation can be expected to be younger and with fewer comorbidities than those hospitalised,[14,19] while those progressing to intensive care can be expected to be older and in worse general health.[3,20] In addition, those being referred to or admitted to intensive care also seem more likely to be male.[3,20] Admission to hospital (and intensive care) is also influenced by a range of supply-side factors, such as availability of beds and criteria for admission, and so the characteristics of those hospitalised do not necessarily only reflect the characteristics accompanying severe illness. These factors, along with geographic variation in populations and transmission dynamics, likely explain some of the heterogeneity seen in those hospitalised with COVID-19 in this study.

### Study limitations

The study was based on routinely-collected data and so, as always, data quality issues must be considered. For instance, individuals were considered as having COVID-19 at time of hospitalisation only if they had a test result or diagnosis indicating the disease, which will have led to the omission of individuals who can be suspected to have had the disease but lacking confirmation of it. Medical conditions may have been underestimated as they were based on the presence of condition codes, with the absence of such a record taken to indicate the absence of a disease. Meanwhile, medication records indicate that an individual was prescribed or dispensed a particular drug, but this does not necessarily mean that an individual took the drug as originally prescribed or dispensed. Our study could be subject to exposure misclassification with false positives if a patient had a medication dispensing event but did not ingest the drug, but may also be subject to false negatives for non-adherent patients who continued their medication beyond the days of supply due to stockpiling. Medication use estimates based on the data collected at the time of hospitalization is particularly sensitive to misclassification, and may conflate baseline concomitant drug history with immediate treatment upon admission. Comparisons of individuals hospitalised with COVID-19 with individuals previously hospitalised with influenza has limitations. In particular, observed differences may be explained by changes in clinical practice or data capture procedures over time, rather than by differences in the individuals themselves. This is likely particularly relevant for comparisons of medication use.

## Conclusion

Rates of comorbidities and medication use are high among individuals hospitalised with COVID-19. Those individuals hospitalised with COVID-19 do not, however, appear to be in worse general health than those typically hospitalised with influenza. Indeed, in many cases, individuals hospitalised with COVID-19 were seen to be younger and healthier than patients hospitalised with seasonal influenza. Patients hospitalised for COVID-19 are also more likely to be male in comparison to those hospitalised with influenza. Protecting those groups known to be vulnerable to influenza is likely to be a useful starting point to minimize the number of hospital admissions needed for COVID-19, but such strategies may need to be broadened so as to reflect the particular characteristics of individuals seen here to have been hospitalised with COVID-19.

## Data Availability

Open Science is a guiding principle within OHDSI. As such, we provide unfettered access to all open-source analysis tools employed in this study via https://github.com/OHDSI/, as well as all data and results artefacts that do not include patient-level health information via http://evidence.ohdsi.org/Covid19CharacterizationHospitalization/.
Data partners contributing to this study remain custodians of their individual patient-level health information and hold either IRB exemption or approval for participation.

## Ethical approvals

All the data partners received Institutional Review Board (IRB) approval or exemption. STARR-OMOP had approval from IRB Panel #8 (RB-53248) registered to Leland Stanford Junior University under the Stanford Human Research Protection Program (HRPP). The use of VA data was reviewed by the Department of Veterans Affairs Central Institutional Review Board (IRB) and was determined to meet the criteria for exemption under Exemption Category 4(3) and approved the request for Waiver of HIPAA Authorization. The research was approved by the Columbia University Institutional Review Board as an OHDSI network study. The IRB number for use of HIRA data was AJIB-MED-EXP-20-065. HM Hospitales and SIDIAP analyses were approved by the Clinical Research Ethics Committee of the IDIAPJGol (project code: 20/070-PCV). The UC-HDC data use was reviewed by Colorado Multi-Institutional Review Board (COMIRB) and was determined to meet the criteria for exemption under Exemption Category 4(3) and approved the request for Waiver of HIPAA Authorization (protocol # 20-0730).

## Acknowledgements

The authors appreciate the Korean Health Insurance Review and Assessment Service for providing data and HM Hospitales for making their data publicly available as part of the COVID Data Save Lives project. UC HDC is supported by the Health Data Compass Data Warehouse project (healthdatacompass.org). VH work was supported by the Intramural Research Program, National Institute of Health.

## Appendix

**Appendix figure A1.**
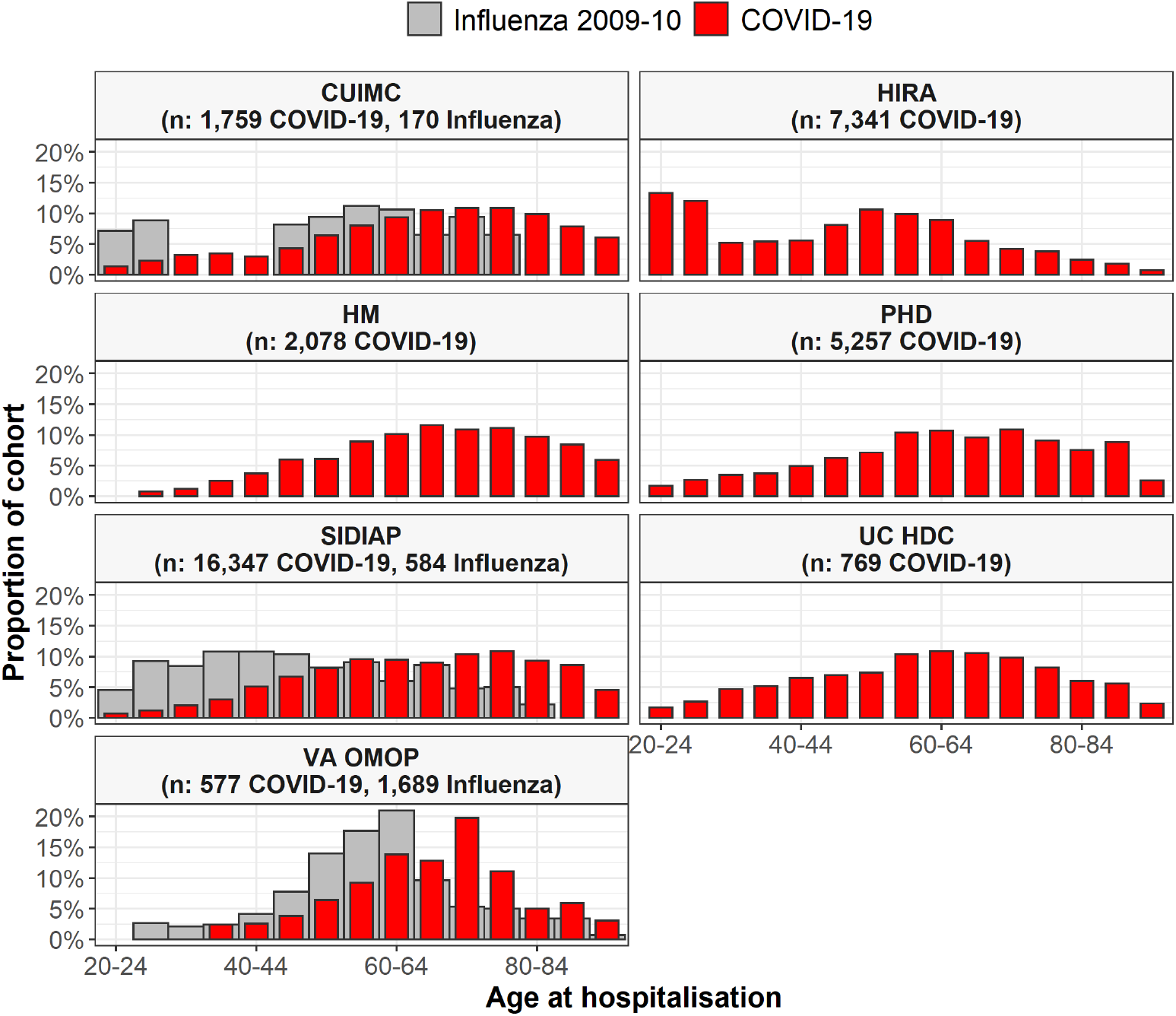
Age of patients hospitalised with COVID-19 and of patients hospitalised with influenza. Individuals hospitalised with COVID-19 between December 2019 and April 2020 compared with those hospitalised with influenza between September 2009 to April 2010. Proportion of cohorts by 5-year age groups, with counts of less than 10 omitted. CUIMC: Columbia University Irving Medical Center; HIRA: Health Insurance Review & Assessment; HM: HM Hospitales; PHD: Premier Healthcare Database; SIDIAP: The Information System for Research in Primary Care; UC HDC: University of Colorado Health Data Compass; VA OMOP: Department of Veterans Affairs.

**Appendix figure A2.**
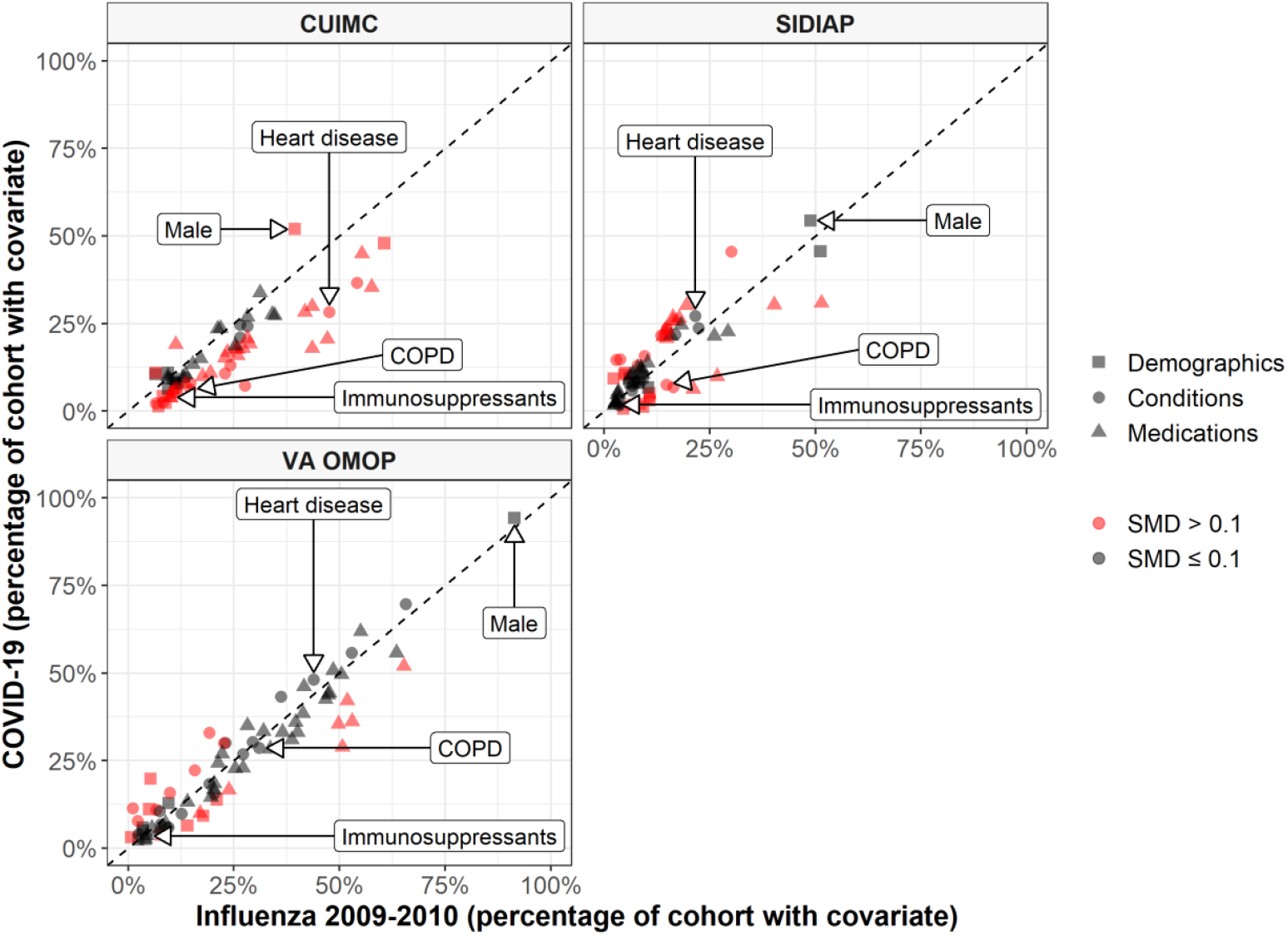
Characteristics of COVID-19 patients compared to 2009-2010 Influenza patients. The plot compares demographics (age and sex), conditions (recorded over the year prior and up to day of hospitalisation), and medications (1) from a year prior to day of hospitalisation, 2) from 30 days prior to day of hospitalisation, and 3) on day of hospitalisation). Each dot represents one of these covariates with the colour indicating the absolute value of the standardised mean difference (SMD), with a SMD above 0.1 taken to indicate a difference in the prevalence of a particular covariate. The proportion male, with heart disease, with chronic obstructive pulmonary disease (COPD) and taking immunosuppressants (over the 30 days prior to hospitalisation) are shown for illustration. CUIMC: Columbia University Irving Medical Center; SIDIAP: The Information System for Research in Primary Care; VA OMOP: Department of Veterans Affairs.

**Appendix table A1.**
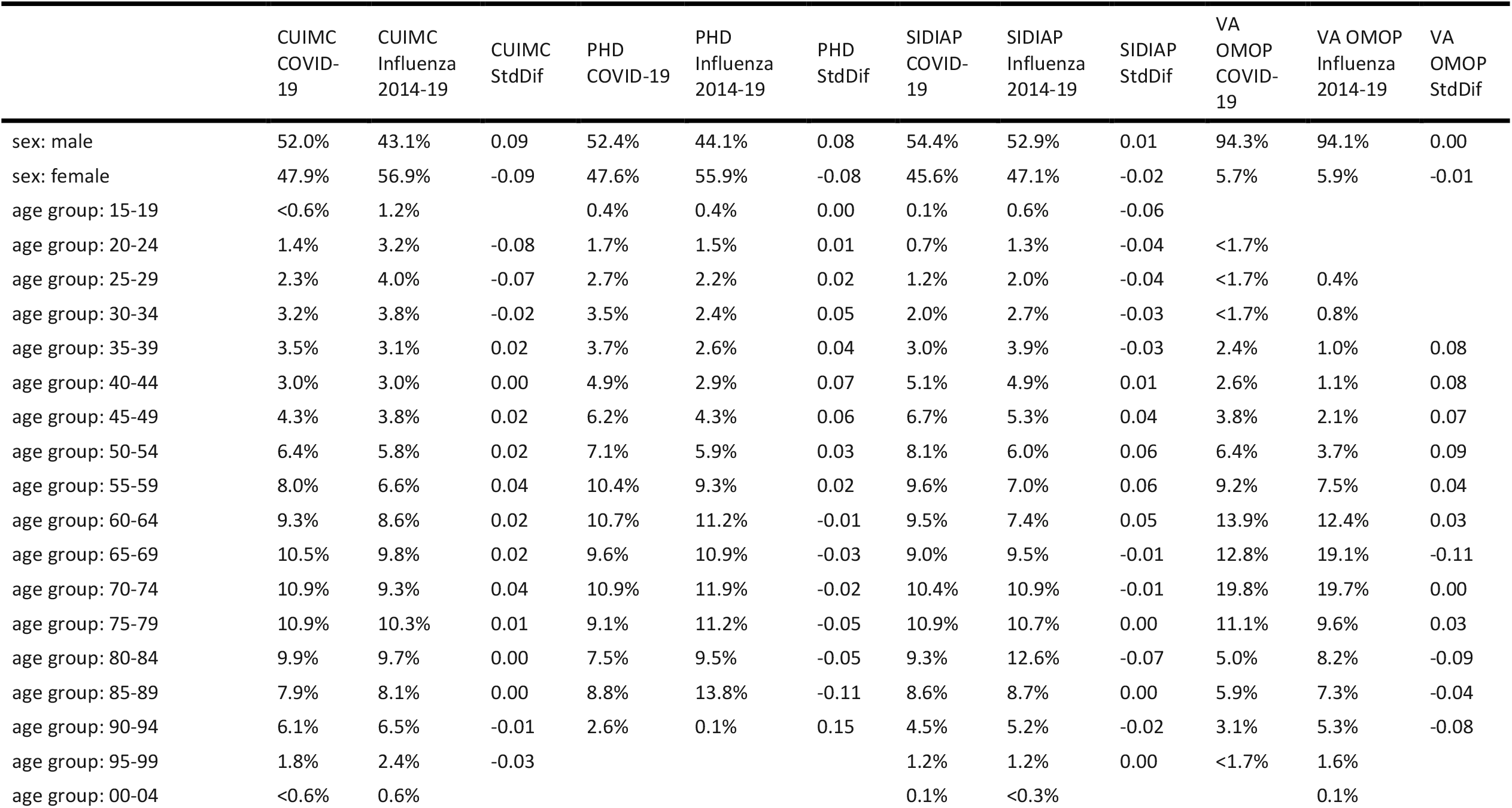
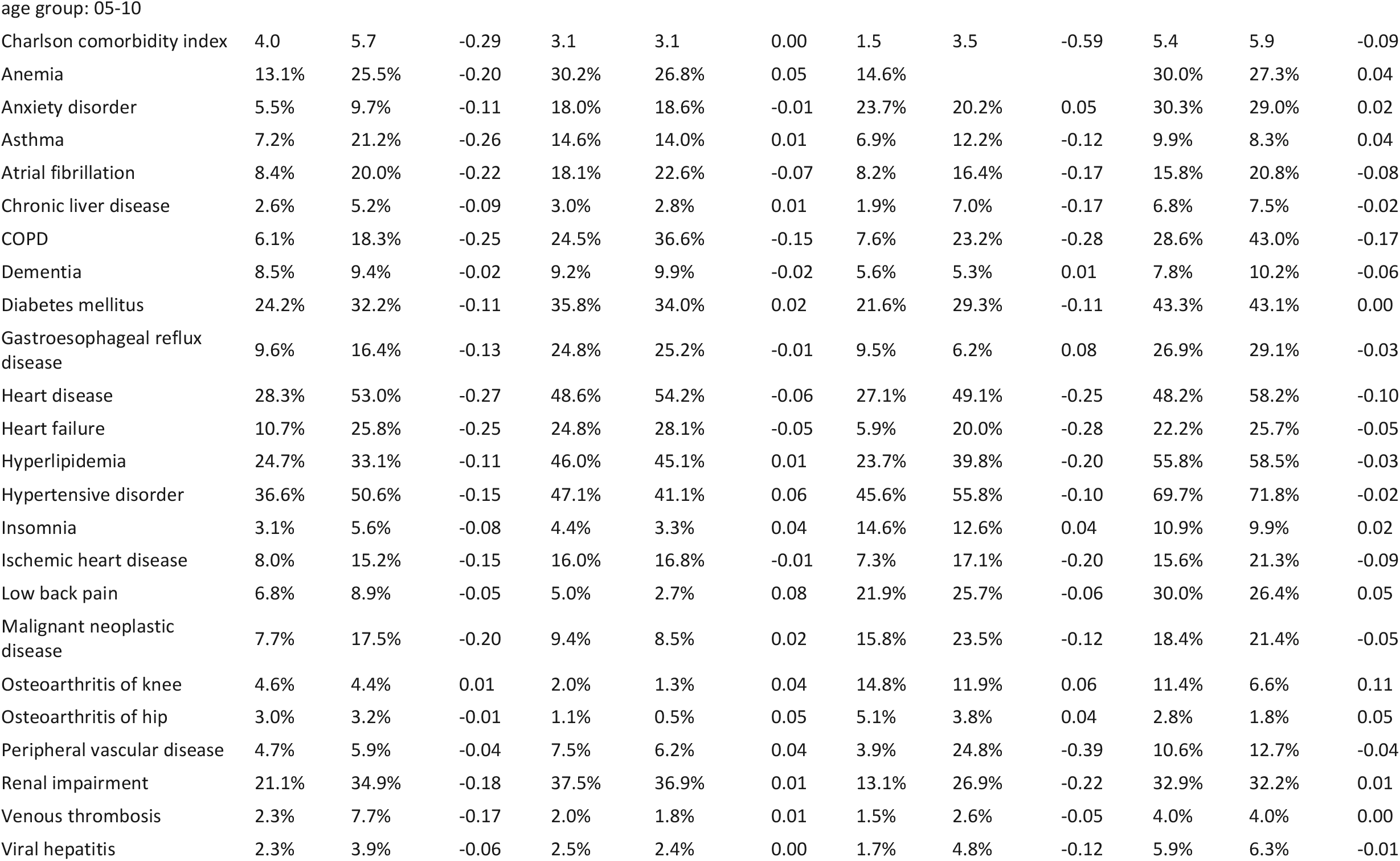
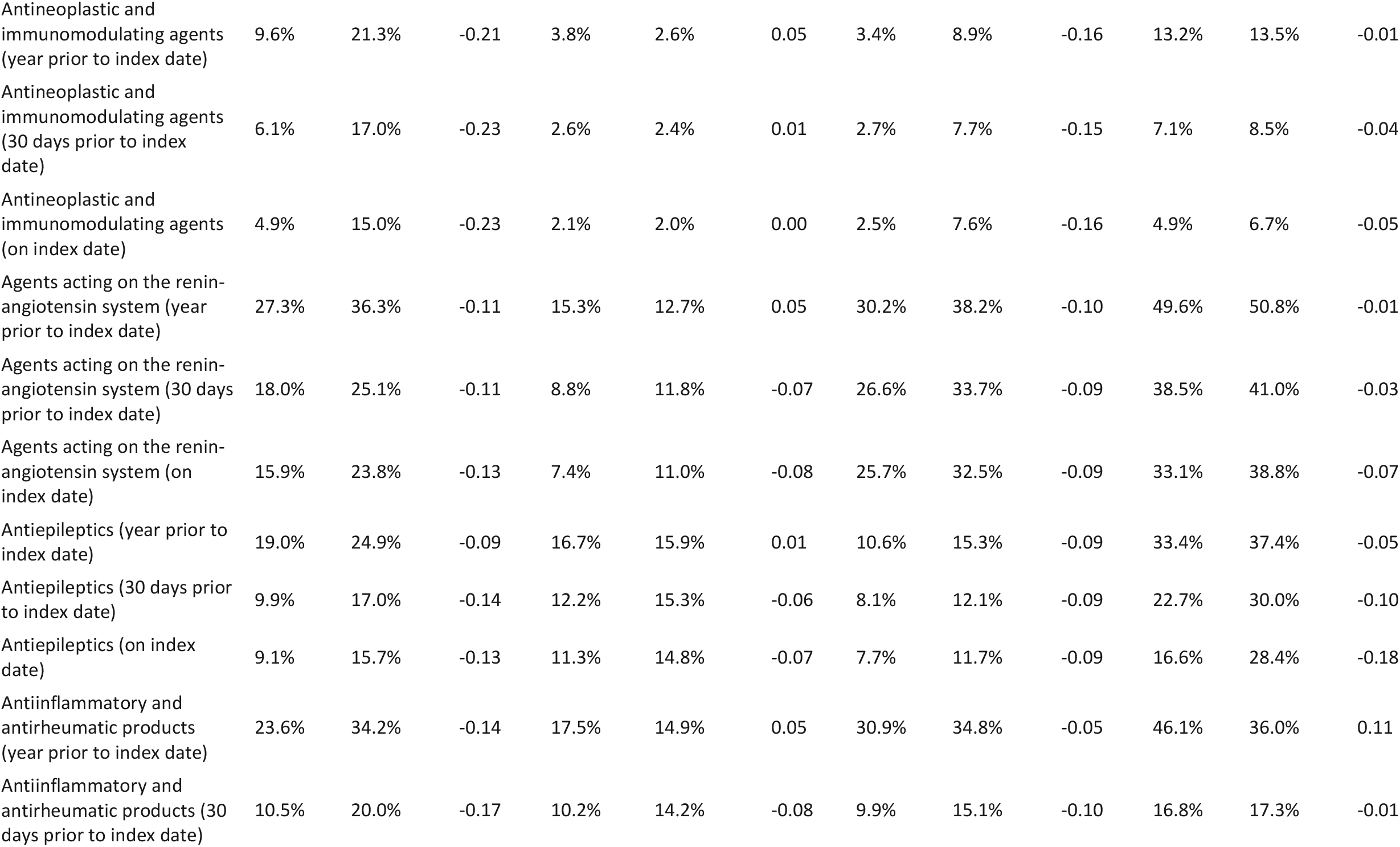
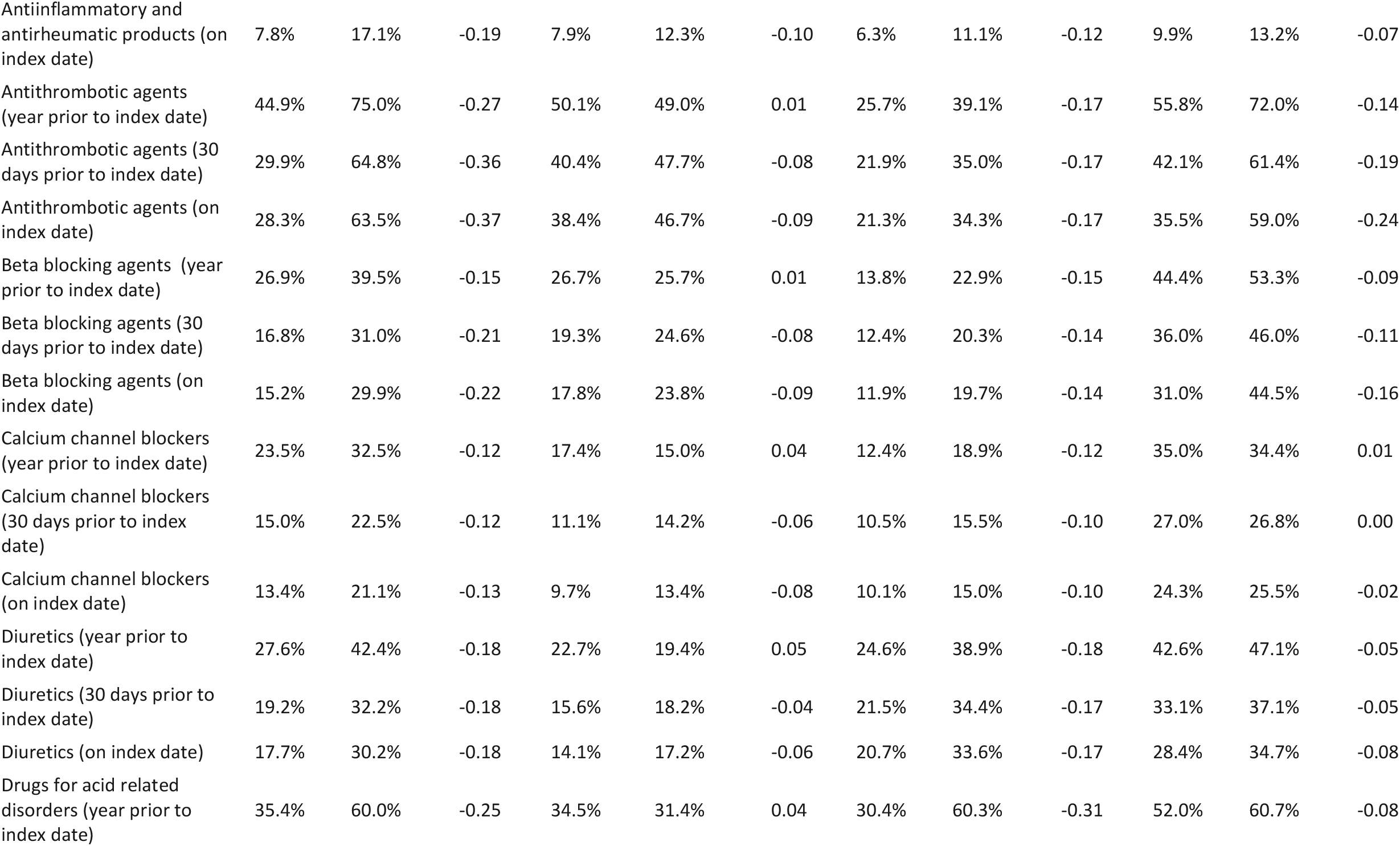
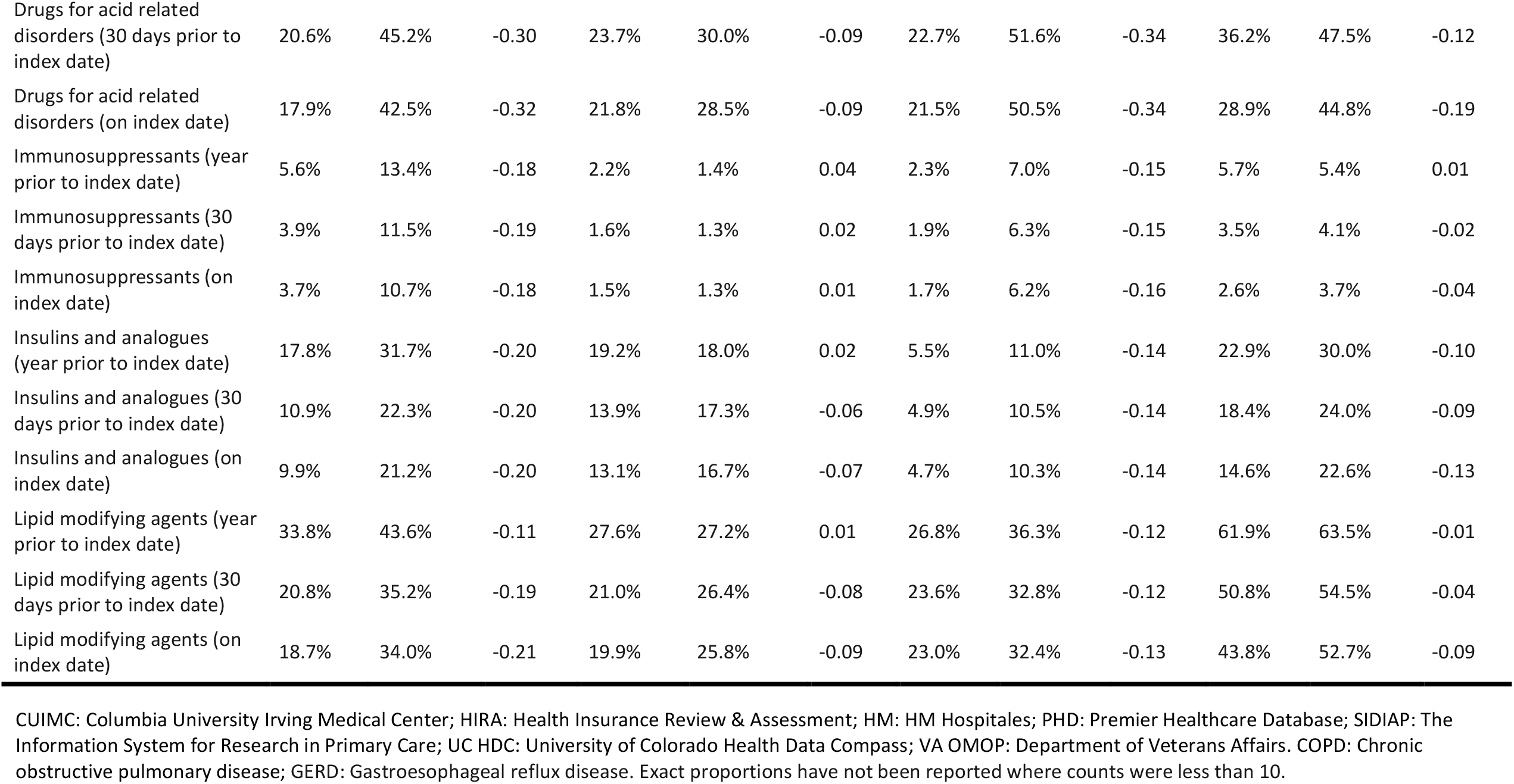
Characteristics of individuals hospitalised with COVID-19 compared with those hospitalised with influenza in 2015-2019

**Appendix table A2.**
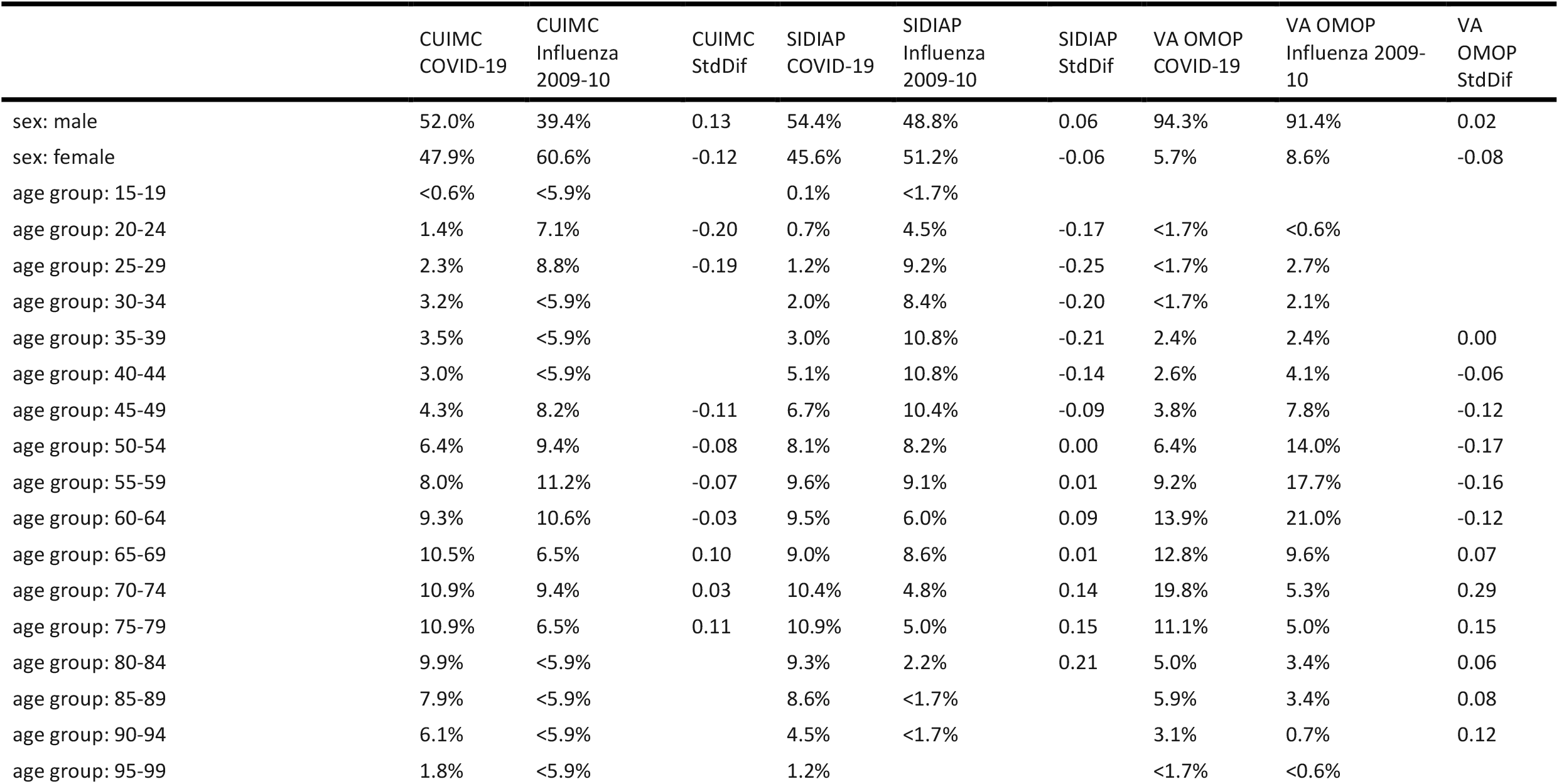
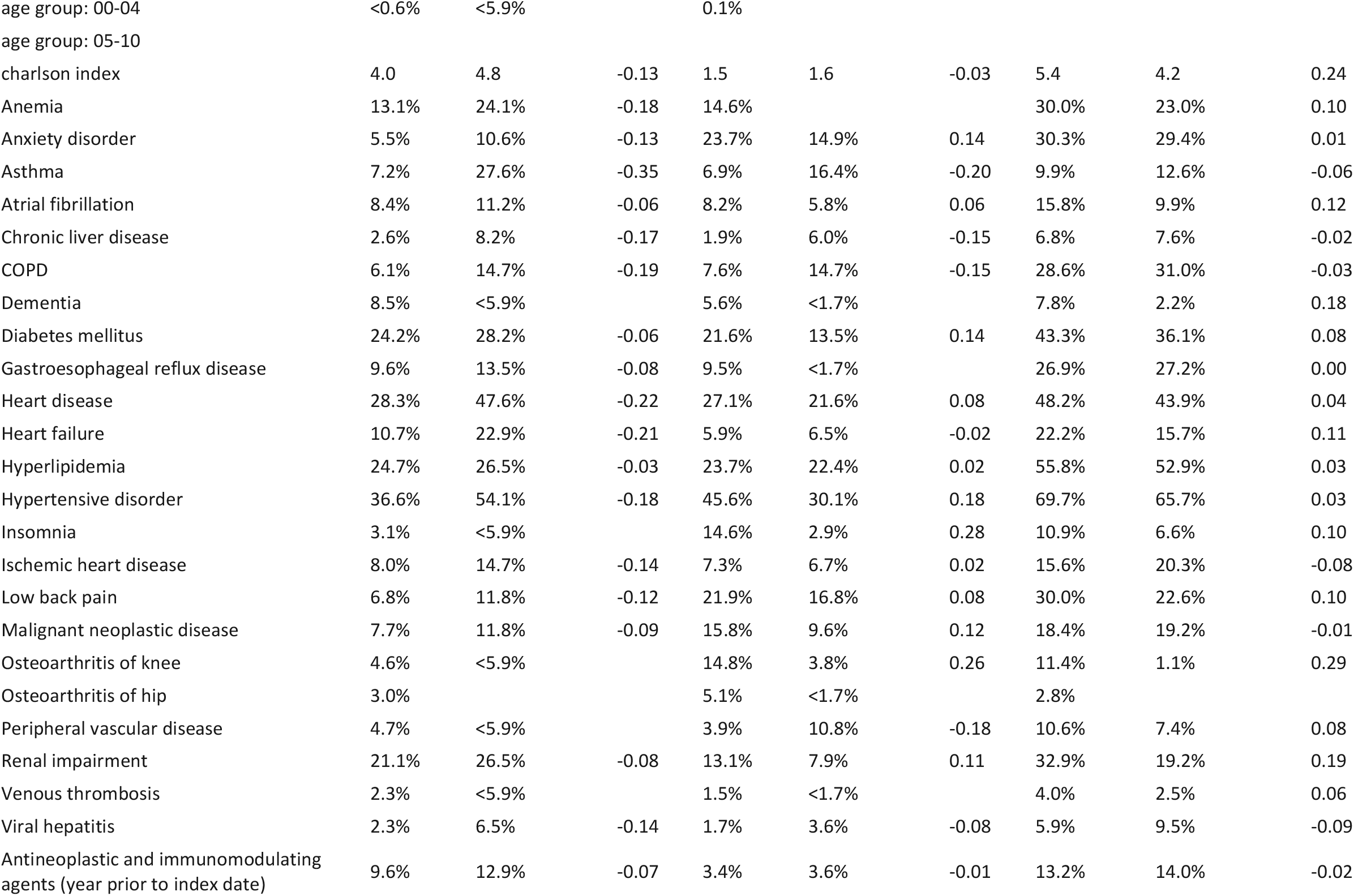
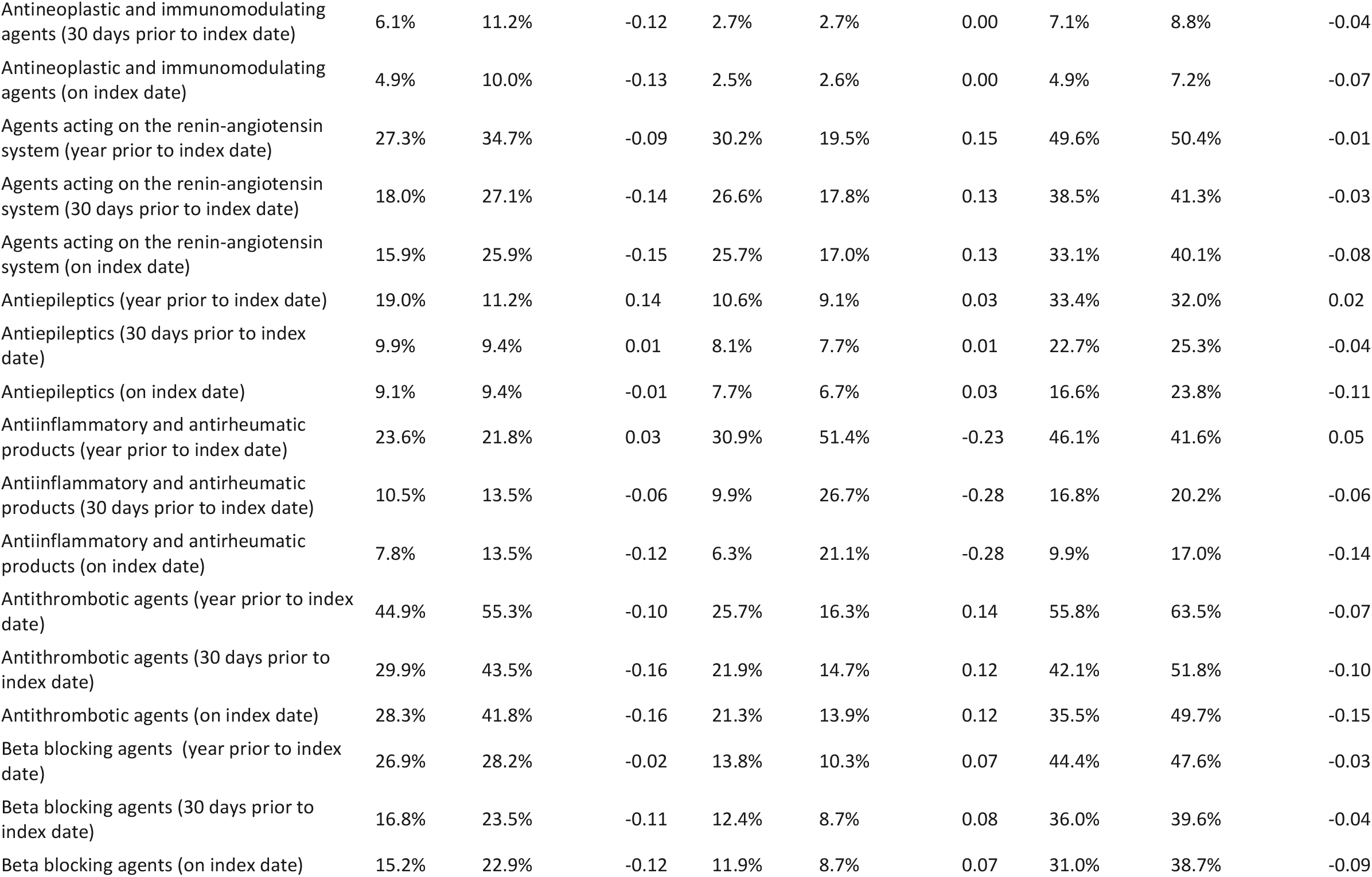
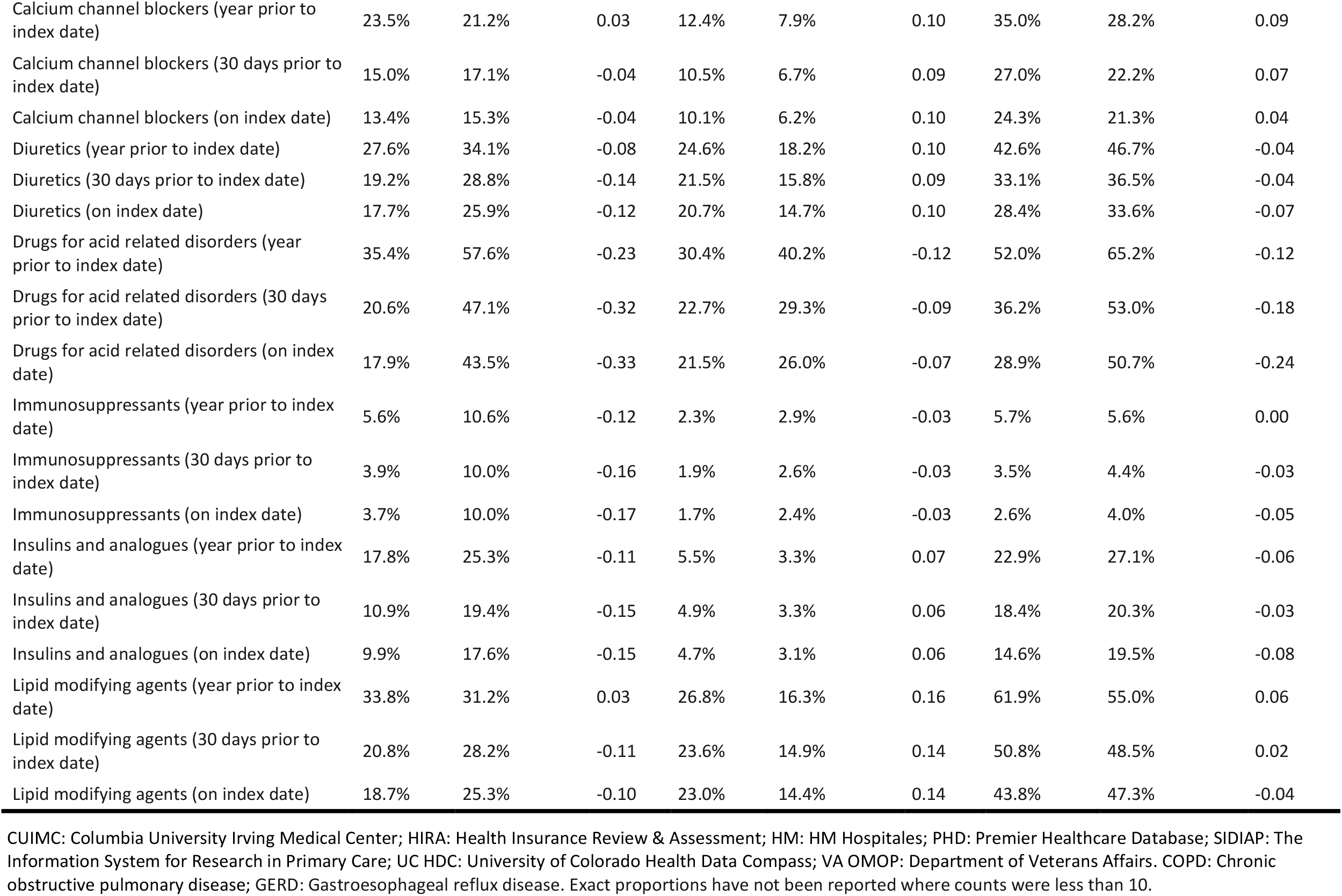
Characteristics of individuals hospitalised with COVID-19 compared with those hospitalised with influenza in 2009-2010

## References

1 Chen N, Zhou M, Dong X, et al. Epidemiological and clinical characteristics of 99 cases of 2019 novel coronavirus pneumonia in Wuhan, China?: a descriptive study. Lancet 2020;395:507–13. doi:10.1016/S0140-6736(20)30211-7

2 Wang D, Hu B, Hu C, et al. Clinical Characteristics of 138 Hospitalized Patients With 2019 Novel Coronavirus–Infected Pneumonia in Wuhan, China. JAMA 2020;323:1061–9. doi:10.1001/jama.2020.1585

3 Bhatraju PK, Ghassemieh BJ, Nichols M, et al. Covid-19 in Critically Ill Patients in the Seattle Region — Case Series. N Engl J Med Published Online First: 30 March 2020. doi:10.1056/NEJMoa2004500

4 Verity R, Okell LC, Dorigatti I, et al. Estimates of the severity of coronavirus disease 2019?: a model-based analysis. Lancet 2020;3099:1–9. doi:10.1016/S1473-3099(20)30243-7

5 Mertz D, Kim TH, Johnstone J, et al. Populations at risk for severe or complicated influenza illness: systematic review and meta-analysis. BMJ Br Med J 2013;347:f5061. doi:10.1136/bmj.f5061

6 Reed C, Chaves SS, Perez A, et al. Complications Among Adults Hospitalized With Influenza: A Comparison of Seasonal Influenza and the 2009 H1N1 Pandemic. Clin Infect Dis 2014;59:166–74. doi:10.1093/cid/ciu285

7 Voss EA, Makadia R, Matcho A, et al. Feasibility and utility of applications of the common data model to multiple, disparate observational health databases. J Am Med Inf Assoc 2015;22:553–64. doi:10.1093/jamia/ocu023

8 Hripcsak G, Duke JD, Shah NH, et al. Observational Health Data Sciences and Informatics (OHDSI): Opportunities for Observational Researchers. Stud Health Technol Inform 2015;216:574–8.http://www.ncbi.nlm.nih.gov/pubmed/26262116 %0A http://www.pubmedcentral.nih.gov/articlerender.fcgi?artid=PMC4815923

9 Garcí;a-Gil MDM, Hermosilla E, Prieto-Alhambra D, et al. Construction and validation of a scoring system for the selection of high-quality data in a Spanish population primary care database (SIDIAP). Inform Prim Care 2011;19:135–45.http://www.ncbi.nlm.nih.gov/pubmed/22688222

10 Kim J-A, Yoon S, Kim L-Y, et al. Towards Actualizing the Value Potential of Korea Health Insurance Review and Assessment (HIRA) Data as a Resource for Health Research: Strengths, Limitations, Applications, and Strategies for Optimal Use of HIRA Data. J Korean Med Sci 2017;32:718–28.https://doi.org/10.3346/jkms.2017.32.5.718

11 Datta S, Posada J, Olson G, et al. A new paradigm for accelerating clinical data science at Stanford Medicine. arXiv 2020.

12 Guan W, Ni Z, Hu Y, et al. Clinical Characteristics of Coronavirus Disease 2019 in China. N Engl J Med Published Online First: 28 February 2020. doi:10.1056/NEJMoa2002032

13 Zhou F, Yu T, Du R, et al. Clinical course and risk factors for mortality of adult inpatients with COVID-19 in Wuhan, China: a retrospective cohort study. Lancet 2020;395:1054–62. doi:10.1016/S0140-6736(20)30566-3

14 Petrilli CM, Jones SA, Yang J, et al. Factors associated with hospitalization and critical illness among 4,103 patients with COVID-19 disease in New York City. medRxiv 2020;:2020.04.08.20057794. doi:10.1101/2020.04.08.20057794

15 Garg S, Kim L, Whitaker M, et al. Hospitalization Rates and Characteristics of Patients Hospitalized with Laboratory-Confirmed Coronavirus Disease 2019 — COVID-NET, 14 States, March 1–30, 2020. MMWR Morb Mortal Wkly Rep Published Online First: 2020. doi:http://dx.doi.org/10.15585/mmwr.mm6915e3

16 Whiting P, Morden A, Tomlinson LA, et al. What are the risks and benefits of temporarily discontinuing medications to prevent acute kidney injury? A systematic review and metaanalysis. BMJ Open 2017;7:e012674. doi:10.1136/bmjopen-2016-012674

17 Horby P, Lim WS, Emberson J, et al. Effect of Dexamethasone in Hospitalized Patients with COVID-19: Preliminary Report. medRxiv 2020;:2020.06.22.20137273. doi:10.1101/2020.06.22.20137273

18 Vogelstein JT, Powell M, Koenecke A, et al. Alpha-1 adrenergic receptor antagonists for preventing acute respiratory distress syndrome and death from cytokine storm syndrome. arXiv 2020.

19 CDC COVID-19 Response Team. Preliminary Estimates of the Prevalence of Selected Underlying Health Conditions Among Patients with Coronavirus Disease 2019 — United States, February 12–March 28, 2020. MMWR Morb Mortal Wkly Rep 2020;69. doi: http://dx.doi.org/10.15585/mmwr.mm6913e2externalicon

20 Grasselli G, Zangrillo A, Zanella A, et al. Baseline Characteristics and Outcomes of 1591 Patients Infected With SARS-CoV-2 Admitted to ICUs of the Lombardy Region, Italy. JAMA Published Online First: 6 April 2020. doi:10.1001/jama.2020.5394

